# Alzheimer’s Disease Classification Confidence of Individuals using Deep Learning on Heterogeneous Data

**DOI:** 10.1101/2024.08.02.24311397

**Authors:** Afolabi Salami Alausa, Jose M. Sanchez-Bornot, Abdoreza Asadpour, Paula L. McClean, KongFatt Wong-Lin, Alzheimer’s Disease Neuroimaging Initiative (ADNI)

## Abstract

Making accurate diagnosis of Alzheimer’s disease (AD) is crucial for effective treatment and management. Although deep learning has been applied to AD classification, it is typically performed at group level, the data used are not sufficiently heterogeneous and comprehensive, and decision confidence is not evaluated at individual (single patient) level. This paper proposed a more practical deep learning approach that not only detects AD stages of individuals, but also provides its corresponding confidence estimation. In particular, in addition to a convolutional neural network (CNN), we incorporated a softmax confidence metric based on the network’s output activity to evaluate its classification confidence. Further, we applied this approach to a heterogeneous and comprehensive data that comprised cognitive and functional assessments, tau-PET and MRI neuroimaging, medical/family history, demographic, and APoE genotype. Importantly, we utilised leave-one-out cross-validation to train the CNN and classify an individual’s healthy control, mild cognitive impairment or AD state, while concurrently estimating each output decision’s confidence. We showed that, over different confidence softmax temperature values, CNN could attain classification accuracies at 83-85% for the three classes while having robust confidence scores of 78-83%. Further improvement in confidence breakdown was achieved using the optimal temperature value in confidence evaluation, with higher confidence scores for correct than error decisions. Overall, the computed classification confidence of an individual may aid clinicians and other stakeholders in understanding the reliability of the model’s decision outcome and offer better trust. The implication of this work may extend to other classification applications, in which the confidence level of a single deep learning-based decision can be evaluated.

## 1 Introduction

Dementia, a prevalent and escalating global health concern, represents a complex spectrum of neurodegenerative disorders that profoundly impact cognitive functions, memory, and daily life activities [1][2]. Alzheimer’s disease (AD) represents the most common cause of dementia [2]. Accurate and early diagnosis of dementia or AD is pivotal for effective therapeutic interventions, management, and improved patient outcomes [3]. However, existing methods for dementia/AD stage classification are often subjective and rely on clinical expertise, making them prone to errors and inconsistencies [4]. The advent of machine learning, including deep learning, has ushered in unprecedented opportunities to harness the potential of heterogeneous data sources, including clinical assessments, neuroimaging, and genetic information, for enhancing the precision of dementia diagnosis and prognosis [4]-[9].

Several machine learning studies have applied deep neural networks or deep learning to dementia diagnosis, due to their high performance [5]-[9]. Most deep learning approaches for dementia/AD (stage) classification have focused on using a single modality such as neuroimaging data due to earlier success in classifying imaging data, or together with few other non-imaging data types [4][7][8][9][10].

Despite their success, these previous studies are limited in their ability to capture more complex and multifaceted nature of dementia/AD, which involves not only changes in brain structure and function, but also cognitive and functional abilities, and other biomarkers and risk factors [4][7][9]. Importantly, classification with deep learning is typically tested across groups rather than on individuals [4]-[9], even though classification of individuals, such as individual patients, is more practical (e.g. [11][12]).

Machine learning also typically computes the confidence of classification outcomes via computing of statistical based confidence intervals over multiple samples [13]-[15], and hardly on individuals. In AD classification, confidence intervals have been used on top of classification or regression (e.g. [12]). Although uncertainty in deep neural networks, such as the various Bayesian and ensemble approaches, have been investigated [13]-[15], there is limited application to AD classification.

In one study [16], the quantification of uncertainty in deep learning-based classification for AD diagnosis was proposed. In this work, it presented a methodology for training the Monte Carlo dropout algorithm [17] by optimising its hyperparameters using Bayesian optimisation [18]. In this way, it improved the quantification of uncertainty in model predictions by giving incorrect AD predictions a high predictive entropy. However, this method was not straightforward, and it is unclear how it can be applied to individual AD diagnosis. Further, it was applied only to imaging data. A more recent study [19] made use of weight perturbation to estimate the variability of predictions by sampling different neural networks similar to the original one at each forward pass. Again, this is a rather cumbersome approach and was applied only to imaging data. Another study, though focused on AD prognosis and not diagnosis, made use of an ensemble-based model where calibrated predictions from multiple pairs (classifier, uncertainty method) are combined to predict whether a cognitive impaired patient will convert to AD [20]. The classifiers used include neural network models. However, this study used five-fold cross-validation (i.e. group-based, unlike [12]), used not comprehensive (only demographic and neuropsychological) data, and did not break down the uncertainty/confidence into correct/error decisions.

In this paper, we propose a deep learning-based approach to classifying an individual’s AD stage using leave-one-out cross validation [12] while applying to a heterogeneous and comprehensive AD data [21] that comprises tau-PET and MRI (regions-of-interest) neuroimaging, medical/family history, cognitive and functional assessments, demographic, and (APoE) genotypes, to embrace the complex and multifaceted nature of AD. Inspired by cognitive neuroscience of decision-making and modelling parsimony, we directly read out the decision confidence score from the network’s output neuronal activity in each classification, providing additional insights into the reliability of our classification. Decision confidence scores are further broken down based on correct and error decisions, and confidence-based temperature is varied to investigate optimality and the robustness of our results.

## 2 Methodology

### 2.1 Data Description

The publicly available Alzheimer’s Disease Neuroimaging Initiative (ADNI) database (adni.loni.usc.edu), particularly the ADNIMERGE-3 open repository, provided the dataset for this investigation. Only ADNI patients who had completed MRI and tau-PET scans (to identify tau deposition) are included in the study. The study measured tau protein deposition for brain imaging using the [^18^F]AV-1451 tracer. Tau protein, a key characteristic marker of AD pathology, is the precise target of this tracer.

The tau and MRI imaging data was combined with baseline study measurements of neuropsychological, medical/family history, and sociodemographic characteristics. There were 224 features in the final dataset: 7 characteristics related to sociodemographic and medical history, 40 scores from cognitive and functional assessments (CFAs), and 177 features from neuroimaging data extracted from tau PET co-registered with MRI data in the form of regions of interest (ROIs) [21]. The data comes with three classes of clinical diagnosis: control normal (CN), AD, and mild cognitive impairment (MCI, which includes prodromal stage of AD).

Sociodemographic data that was gathered included age, gender, years of schooling, number of copies of APoE ε4 gene variant (APoE4), and whether AD ran in the family on both the mother’s and father’s lines. It is established that the APoE4 variant is linked to a higher chance of AD, we encoded the APoE4 data as 0, 1, or 2 for easier subsequent analysis. CFAs were also included in the data, comprising Clinical Dementia Rating (CDR), Mini-Mental State Exam (MMSE), Alzheimer’s Disease Assessment Scale (ADAS), Neuropsychological Inventory (NPI), Geriatric Depression Scale (GDS), Cognitive Battery Assessment, Modified Hachinski Ischemia Scale, Neuropsychological Battery Test, logical memory immediate recall test (LMIT), and logical memory delayed recall test (LMDT). Other CFAs were excluded due to large proportion of missing data [21]. Further, we used individual subscales from the NPI and total scores as well as individual question scores from the ADAS, providing a higher level of CFA granularity [21].

Ultimately, the dataset contained information on 559 participants following the combination and pre-processing of all the obtained data. Within this, 363 participants were clinically diagnosed as control normal (CN), 137 had MCI, and 59 had AD, and these AD stages were used as targets or class labels for model training. The dataset in this study was the same as in our previous study [21].

### 2.2 Data Preparation and Preprocessing

As MCI and AD classes were substantially smaller in size than CN, there could be bias in the models in terms of poorer model performance for the minority class, as the learning algorithm might be swayed by the majority class CN. To mitigate any imbalances in the classes, the synthetic minority over-sampling technique (SMOTE) [22] was employed. SMOTE creates synthetic samples for minority classes by interpolating between existing ones while preserving the minority class’s characteristics, thereby balancing the class distribution. After applying SMOTE, all three classes were balanced, and we had a total of 1089 samples, i.e. 363 samples per class.

### 2.3 Model Description

For classification, we used the convolutional neural network (CNN) given its known superior performance in AD classification (e.g. [6]). CNN was used due to its superior performance as compared to traditional machine learning methods [4]-[10], yet not overly complicated (e.g. compared to recurrent neural networks) for applications. Following data balancing, the CNN model architecture is defined using the Keras Sequential API. The architecture consisted of three dense layers: the first two layers have rectified linear unit (ReLU) activation functions for introducing nonlinearity, while sigmoidal function was used in the third, output layer.

Following these layers, and inspired by neuroscience [23] and model parsimony, a custom layer (called ConfidenceSoftmax) with softmax function and temperature scaling was implemented to the output and computed the confidence score for each model prediction. The softmax function *σ* for prediction of class *i* is described by

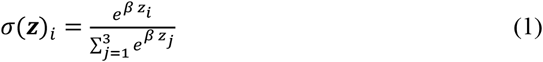

where *β* = 1/*T*, the reciprocal of the temperature *T*, and the summation is over the three output neuronal activities *z*_*j*_ for class *j* = 1,2,3. As *T* approaches 0, the function becomes highly confident (i.e., the predicted probabilities become more extreme). Conversely, as *T* is large, the predicted probabilities become more uniform and thus has low confidence. *T* values from 0.1 to 10 were explored. Fine-tuning of *T* involves selecting a value that optimally balances the confidence and accuracy of the model’s predictions using the validation dataset.

The input layer had 223 neurons, which equate to the number of input variables (i.e. number of columns from the dataset sans participant identification number). The second layer, i.e. the first hidden layer, and the next hidden layer each had 64 neurons. These neuron numbers were tuned to maximise classification performance. Then, the following layer, the output layer, had 3 neurons for the 3-class (CN, MCI and AD) classification, which were then connected to the confidence evaluation layer. A schematic of the neural network architecture is shown in Fig. 1A.

**Fig. 1.**
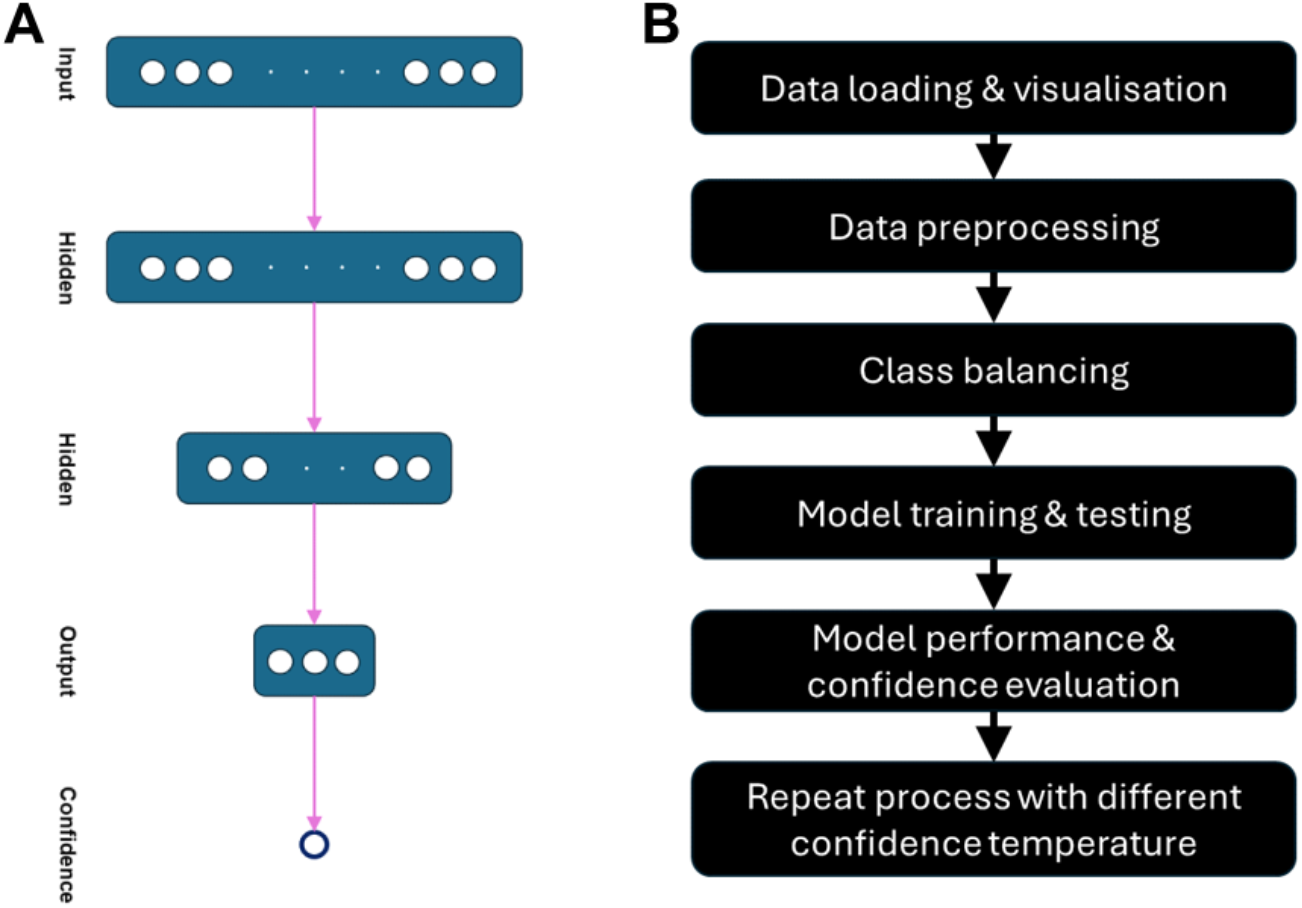
Overall modelling and process. (A) Schematic diagram of the convolutional neural network model architecture. (B) Summary of the overall process.

### 2.4 Model Training, Testing and Evaluation

To test for an individual (single sample), the model was trained and evaluated using leave-one-out cross-validation (LOOCV) [24][12]. Within each iteration of LOOCV, the entire dataset was split into training and testing datasets, with a single data point held out for testing. A neural network model with a specified temperature (e.g., of 1.5 value) was created for model training using 50 epochs and 32 batch size. Each epoch represents one complete pass through the entire training dataset. A validation set was used to monitor the model’s performance and prevent overfitting. The validation loss and accuracy were recorded at each epoch. After initial training, temperature scaling was applied to the logits (output before the softmax layer) to fine-tune the confidence of predictions.

Within each training iteration, the Adam optimiser was used, a popular optimizer known for its efficiency in updating weights during the training process [25]. The standard sparse categorical cross-entropy loss function was used to measure the discrepancy between the predicted class probabilities and the true labels. This loss function is well-suited for multi-class classification tasks with integer-encoded labels. The model exhibited signs of overfitting during the initial epochs, indicated by a significant gap between training and validation loss. This was addressed by implementing dropout layers and early stopping to halt training when validation loss did not improve over several epochs.

The trained model then made predictions on the single held-out test sample in the testing dataset. Classification accuracy was calculated, and the results included predicted class, true class, accuracy, and confidence score for each iteration. To evaluate the classification performance, the average accuracy, F1 score, precision and recall were used. Stability of the network’s final outputs (e.g. accuracy and confidence) was also checked by repeating the whole process 5 times. A summary of the overall process is illustrated in Fig. 1B.

### 2.5 Software and Hardware

The model architecture was defined using the Keras Sequential API. The computations were performed using Python within the Google Colab environment, and executed on a machine with a base speed of 2.40 GHz, equipped with an Intel(R) HD Graphics 520 GPU. Code repository is available at https://github.com/Afolabialausa/Alzheimer-s-Disease-Classification-.git The original ADNI dataset was not included as part of this repository. Requests to access the original datasets should be directed to ADNI (http://adni.loni.usc.edu/).

## 3 Results

### 3.1 Model accuracy and confidence for individual prediction

We began our investigation by first setting the temperature of the confidence softmax function to be 1.0. We found the average accuracy of the 3-class classification to be 0.8421. For completeness, we also evaluated the F1 score, precision and recall and they were 0.8423, 0.8425 and 0.8421, respectively. Although the classification accuracies seemed relatively high and consistent, the standard deviation e.g. for the average accuracy was 0.3647, which was expected given the conservative LOOCV approach applied, i.e. there is relatively higher variation for testing single individual sample or participant. In comparison, the average confidence score was 0.8066, which was reasonable for high average accuracy. Yet, the standard deviation of the confidence score was 0.1681, which was smaller than that of the average accuracy of 0.3647.

Next, we delved deeper into the comparison between the classification performance and confidence score. Fig. 2 shows a confusion matrix for the 3-class classification. Consistent with the high average accuracy, we could observe that the diagonal values of correctly predicted classes were high (CN: 303 cases; MCI: 285 cases; and AD: 329 cases). The off-diagonal misclassification cases were high only for CN-MCI cases, which was not surprising given the mixed types of cases for MCI (some might not lead to AD [4]). For the CN-AD off-diagonal cases, there were only a handful of misclassified cases. By aggregating the CN, MCI and AD classes, we also computed similar confidence scores for the true positive (TP), true negative (TN), false positive (FP), and false negative (FN) cases. We found the confidence scores for TP, TN, FP and FN to be 0.6723, 0.7511, 0.6538 and 0.7796, respectively. Ideally, we would prefer the correctly classified (TP and TN) cases to have higher confidence scores than that of incorrectly classified (FP and FN) cases. However, we found the confidence scores of the FP and FN cases to be similar to those of TP and TN. This suggested overconfidence of errors in the model.

**Fig. 2.**
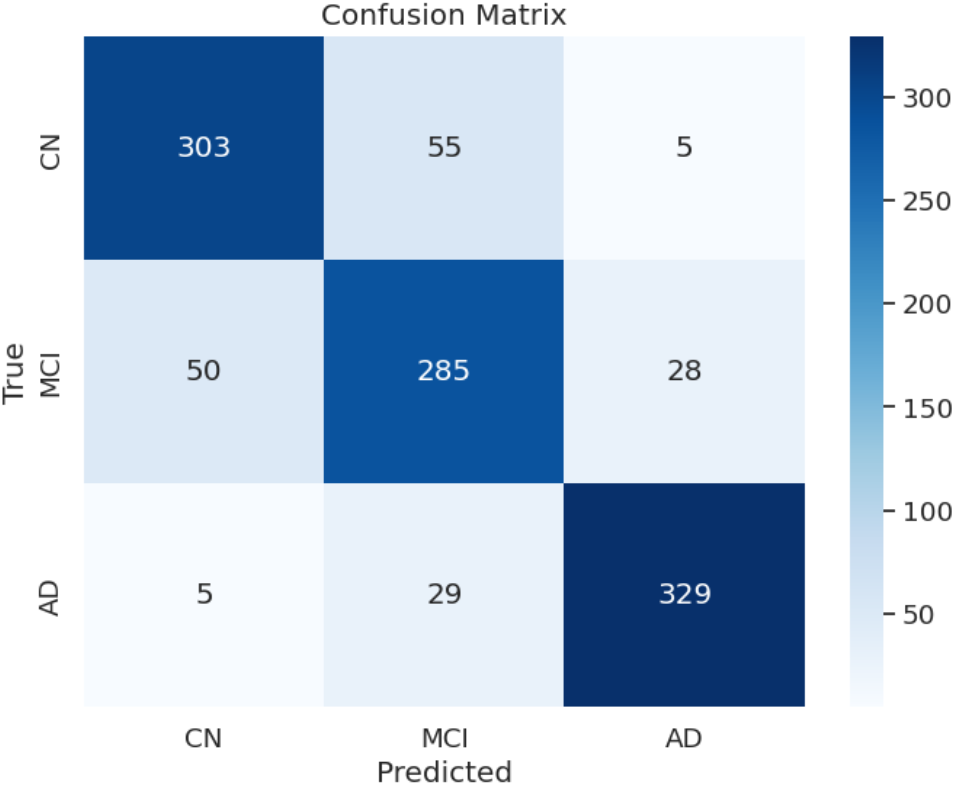
Confusion matrix for the 3-class classification of the class-balanced data using temperature of 1.0. CN: control normal; MCI: mild cognitive impairment; AD: Alzheimer’s disease. Colour bar: case number.

To resolve this overconfidence issue, we next investigated the temperature values of the confidence function from 0.5 (more conservative) to 2.5 (more exploratory) by repeating the above analytical process. Table 1 gives a summary of the results. First, we observed that the model’s classification accuracy (average accuracy, F1 score, precision and recall) performances were robust across temperature values. Second, we observed that the average confidence scores were also similar across temperature values. These provides reassurance to the model’s function. Importantly, we found that the confidence scores for the TP, TN, FP and FN cases could vary substantially as we vary the temperature values. A low temperature of 0.5 led to overconfidence (>0.7) in all four cases, while a high temperature of 2.5 led to underconfidence (<0.7). The optimal temperature for our study was found to have a value of 2.0, as the TP and TN cases had higher confidence scores than those of the FP and FN cases. Further, the standard deviation of the confidence score was also noticeably lower than with other temperature values.

**Table 1.** Summary of model performance and confidence over different confidence temperatures. SD: standard deviation. Bold: optimal set of confidence scores. Shaded: for confidence.

| Confidence temperature | 0.5 | 1.0 | 1.5 | 2.0 | 2.5 |
| --- | --- | --- | --- | --- | --- |
| Average accuracy | 0.8448 | 0.8421 | 0.8320 | 0.8466 | 0.8476 |
| SD accuracy | 0.3621 | 0.3647 | 0.3739 | 0.3603 | 0.3594 |
| F1 score | 0.8449 | 0.8423 | 0.8314 | 0.8467 | 0.8472 |
| Precision | 0.8456 | 0.8425 | 0.8314 | 0.8469 | 0.8470 |
| Recall | 0.8448 | 0.8421 | 0.8320 | 0.8466 | 0.8476 |
| Average confidence | 0.8152 | 0.8066 | 0.7888 | **0.8188** | 0.8163 |
| SD confidence | 0.1628 | 0.1681 | 0.1678 | **0.1578** | 0.1613 |
| TP confidence | 0.8024 | 0.6723 | 0.9401 | **0.8415** | 0.6889 |
| TN confidence | 0.8152 | 0.7511 | 0.5369 | **0.7839** | 0.6621 |
| FP confidence | 0.8084 | 0.6538 | 0.6690 | **0.6946** | 0.6871 |
| FN confidence | 0.7288 | 0.7796 | 0.7783 | **0.7238** | 0.6297 |

## 4 Discussion

In this study, we have implemented a deep learning based approach on a heterogeneous and comprehensive AD dataset for individualised AD stage classification and confidence estimation. Using the leave-one-out cross validation approach and a softmax function, we could evaluate the confidence for every model output, i.e. the model’s decision, of an individual. Our results showed that the model attained high classification accuracy for the three classes of CN, MCI and AD, despite high heterogeneity in the data. We showed that varying the confidence’s softmax temperature parameter did not affect the average confidence scores. However, to be more in line with the cognitive (neuro)science of decision-making [23][26] and hence gain better user’s trust, the right temperature was required to attaining higher confidence scores for correctly classified (TP and TN) cases than that of incorrectly classified (FP and FN) cases. In this specific dataset and model, the confidence temperature was found to be at 2.0. Overall, temperature scaling effectively calibrates the model’s confidence, impacting its performance and reliability.

While previous approaches have made significant strides in using deep learning for dementia diagnosis [4]-[10], they often focused on single data types, lacked individual prediction confidence, and provided group-level confidence intervals. The proposed approach overcome these limitations by introducing a neuro-inspired deep learning model that simultaneously classifies dementia stages and reports confidence levels for each prediction, evaluated within a LOOCV framework for individual case handling. This enhances the model’s practical utility and reliability in real-world clinical settings. While our deep learning approach shows promise for individualised AD stage classification and confidence estimation, the current study has some notable limitations. First, the deep network achieves a lower 3-class classification accuracy (83-85%) than that of our previous study (88-90%) using graph neural network, another form of deep learning, on the same dataset [21]. A direction to improve the classification accuracy could be to perform feature selection prior to classification [12][21][27]. Feature selection may also enhance the TP/TN-vs-FP/FN confidence score difference. However, we could not directly compare the results in this previous study [21] as the cross-validation approach was different. In particular, in the current study, LOOCV was used, which is a highly conservative approach with expected lower classification accuracy. Future work will more directly compare our current model with other classification models endowed with confidence or uncertainty evaluation [13][14][15].

Future work should also investigate whether the model in the current study can be generalised to other dementia or AD datasets, including well curated open datasets or more messy clinical datasets [4][28]. In particular, the data used in the current study had large number of features with respect to the number of samples, which may potentially lead to model overfitting. Datasets with smaller number of features or larger sample sizes should be explored.

Although we have advanced a step towards better interpretability and trust in the model, further steps are needed for facilitating clinical decision-making and engendering trust among healthcare practitioners [29]. Future work can perhaps address this by involving clinicians’ insights and opinions as part of the model development – humans-in-the-loop approach [28][30]. Further, LOOCV may be prone to overfitting while being computationally expensive, and future work can evaluate how real-time clinical decisions can be better achieved using this approach. Addressing these limitations requires ongoing efforts to refine preprocessing techniques, explore alternative validation approaches, and enhance the interpretability of confidence measures. Ensuring the model’s robustness and applicability in real-world clinical settings will depend on overcoming these challenges and validating the approach across diverse and representative datasets.

In conclusion, we have demonstrated the feasibility of using confidence evaluation in deep network model for detecting AD stage of an individual. The use of confidence score for individual diagnosis offers clinicians and other stakeholders valuable insights into the reliability of the model predictions, thereby aiding in informed decision-making, while enhancing transparency and trust in the diagnostic process. Accurate classification with associated confidence scores in clinical decision support systems can also inform and reduce the likelihood of misdiagnosis. This is crucial for timely and appropriate management treatment and intervention and hence improve patient outcomes.

## Data Availability

Code repository is available at https://github.com/Afolabialausa/Alzheimer-s-Disease-Classification-.git The original ADNI dataset was not included as part of this reposi-tory. Requests to access the original datasets should be directed to ADNI (http://adni.loni.usc.edu/).

## Acknowledgement

We thank Marinus Toman for helpful comments and suggestions. Data collection and sharing for this project was funded by the Alzheimer’s Disease Neuroimaging Initiative (ADNI) (National Institutes of Health Grant U01 AG024904) and DOD ADNI (Department of Defense award number W81XWH-12-2-0012). ADNI is funded by the National Institute on Aging, the National Institute of Biomedical Imaging and Bioengineering, and through generous contributions from the following: AbbVie, Alzheimer’s Association; Alzheimer’s Drug Discovery Foundation; Araclon Biotech; BioClinica, Inc.; Biogen; Bristol-Myers Squibb Company; CereSpir, Inc.; Cogstate; Eisai Inc.; Elan Pharmaceuticals, Inc.; Eli Lilly and Company; EuroImmun; F. Hoffmann-La Roche Ltd and its affiliated company Genentech, Inc.; Fujirebio; GE Healthcare; IXICO Ltd.; Janssen Alzheimer Immunotherapy Research & Development, LLC.; Johnson & Johnson Pharmaceutical Research & Development LLC.; Lumosity; Lundbeck; Merck & Co., Inc.; Meso Scale Diagnostics, LLC.; NeuroRx Research; Neurotrack Technologies; Novartis Pharmaceuticals Corporation; Pfizer Inc.; Piramal Imaging; Servier; Takeda Pharmaceutical Company; and Transition Therapeutics. The Canadian Institutes of Health Research is providing funds to support ADNI clinical sites in Canada. Private sector contributions are facilitated by the Foundation for the National Institutes of Health (www.fnih.org). The grantee organization is the Northern California Institute for Research and Education, and the study is coordinated by the Alzheimer’s Therapeutic Research Institute at the University of Southern California. ADNI data are disseminated by the Laboratory for Neuro Imaging at the University of Southern California.

## References

1. 2024 Alzheimer’s disease facts and figures. Alzheimer’s & Dementia 20(5), 3708–3821 (2024). doi: 10.1002/alz.13809.

2. Gale, S.A., Acar, D., Daffner, K.R.: Dementia. The American Journal of Medicine 131(10), 1161–1169 (2018).

3. Alzheimer’s Disease International. World Alzheimer Report 2011: The benefits of early diagnosis and intervention. London: ADI; 2011. https://www.alz.co.uk/research/WorldAlz-heimerReport2011.pdf, xlast accessed 2024/06/02.

4. Wong-Lin, K., McClean, P.L., McCombe, N., Kaur, D., Sanchez-Bornot, J.M., Gillespie, P., Todd, S., Finn, D.P., Joshi, A., Kane, J., McGuinniess, B.: Shaping a data-driven era in dementia care pathway through computational neurology approaches. BMC Medicine 18(1), 398 (2020). doi: 10.1186/s12916-020-01841-1.

5. Khojaste-Sarakhsi M., Haghighi, S.S., Ghomi, S.M.T.F., Marchiori, E.: Deep learning for Alzheimer’s disease diagnosis: A survey. Artificial Intelligence in Medicine 130, 102332 (2022). doi: 10.1016/j.artmed.2022.102332.

6. Fathi, S., Ahmadi, M., Dehnad, A.: Early diagnosis of Alzheimer’s disease based on deep learning: A systematic review. Computers in Biology and Medicine 146, 105634 (2022). doi: 10.1016/j.compbiomed.2022.105634.

7. Tsang, G., Xie, X., Zhou, S.M.: Harnessing the power of machine learning in dementia informatics research: Issues, opportunities, and challenges. IEEE Reviews in Biomedical Engineering 13, 113–129 (2020). doi: 10.1109/RBME.2019.2904488.

8. Ahmed, M.R., Zhang, Y., Feng, Z., Lo, B., Inan, O.T., Liao, H.: Neuroimaging and machine learning for dementia diagnosis: Recent advancements and future prospects. IEEE Reviews in. Biomedical Engineering 12, 19–33 (2019). doi: 10.1109/RBME.2018.2886237.

9. Javeed, A., Dallora, A.L., Berglund, J.S., Ali, A., Ali, L., Anderberg, P.: Machine learning for dementia prediction: A systematic review and future research directions. Journal of Medical Systems 47, 17 (2023). doi: 10.1007/s10916-023-01906-7.

10. Qiu, S., Joshi, P.S., Miller, M.I., Xue, C., Zhou, X., Karjadi, C., et al.: Development and validation of an interpretable deep learning framework for Alzheimer’s disease classification. Brain 143(6), 1920–1933 (2020).

11. Youssofzadeh, V., McGuinness, B., Maguire, L.P., Wong-Lin, K.: Multi-kernel learning with Dartel improves combined MRI-PET classification of Alzheimer’s disease in AIBL data: Group and individual analyses. Frontiers in Human Neuroscience 11, 380 (2017).

12. Bucholc, M., Ding, X., Wang, H., Glass, D.H., Wang, H., Prasad, G., Maguire, L.P., Bjourson, A.J., McClean, P.L., Todd, S., Finn, D.P., Wong-Lin, K.: A practical computerized decision support system for predicting the severity of Alzheimer’s disease of an individual. Expert Systems with Applications 130, 157–171 (2019).

13. Abdar, M., Pourpanah, F., Hussain, S., Rezazadegan, D., Liu, L., Ghavamzadeh, M., Fieguth, P., Cao, X., Khosravi, A., Acharya, U.R., Makarenkov, V., Nahavandi, S.: A review of uncertainty quantification in deep learning: Techniques, applications and challenges. Information Fusion 76, 243–297 (2021).

14. Gawlikowski, J., Tassi, C.R.N., Ali, M., Lee, J., Humt, M., Feng, J., Kruspe, A., Triebel, R., Jung, P., Roscher, R., Shahzad, M., Yang, W., Bamler, R., Zhu, X.Y.: A survey of uncertainty in deep neural networks. Artificial Intelligence Review 56(1), 1513–1589 (2023). doi: 10.1007/s10462-023-10562-9.

15. Nemani, V., Biggio, L., Huan, X., Hu, Z., Fink, O., Tran, A., Wang, Y., Zhang, X., Hu, C.: Uncertainty quantification in machine learning for engineering design and health prognostics: A tutorial. Mechanical Systems and Signal Processing 205, 110796, (2023). doi: 10.1016/j.ymssp.2023.110796.

16. Roshanzamir, M., Shamsi, A., Asgharnezhad, H., Alizadehsani, R., Hussain, S., Moosaei, H., Mohammadi, A., Acharva, U.R., Alinejad, H.: Quantifying uncertainty in automated detection of Alzheimer’s patients using deep neural network. (2023) doi: 10.20944/pre-prints202301.0148.v1.

17. Srivastava, N., Hinton, G., Krizhevsky, A., Sutskever, I., Salakhutdinov, R.: Dropout: A simple way to prevent neural networks from overfitting. Journal of Machine Learning Research 15, 1929–1958 (2014).

18. Frazier, P.I.: A tutorial on Bayesian optimization. 1807.02811.

19. Ferrante, M., Boccato, T., Toschi, N.: Enabling uncertainty estimation in neural networks through weight perturbation for improved Alzheimer’s disease classification. Frontiers in Neuroinformatics 18:1346723 (2024). doi: 10.3389/fninf.2024.1346723.

20. Pereira, T., Cardoso, S., Guerreiro, M., Mendonça, A.de, Madeira, S.C.; Alzheimer’s Disease Neuroimaging Initiative: Targeting the uncertainty of predictions at patient-level using an ensemble of classifiers coupled with calibration methods, Venn-ABERS, and Conformal Predictors: A case study in AD. Journal of Biomed Informatics 101, 103350 (2020). doi: 10.1016/j.jbi.2019.103350.

21. McCombe, N., Bamrah, J., Sanchez-Bornot, J.M., Finn, D.P., McClean, P.L., Wong-Lin, K.: Alzheimer’s disease classification using cluster-based labelling for graph neural network on heterogeneous data. Healthcare Technology Letters 9(6), 102–109 (2022).

22. Chawla, N.V., Bowyer, K.W., Hall, L.O., Kegelmeyer, W.P.: SMOTE: Synthetic minority over-sampling technique. Journal of Artificial Intelligence Research 16(1), 321–357 (2002).

23. Kiani, R., Shadlen, M.N.: Representation of confidence associated with a decision by neurons in the parietal cortex. Science 324(5928), 759–764 (2009).

24. Stone, M.: Cross-validatory choice and assessment of statistical predictions. Journal of the Royal Statistical Society, Series B (Methodological) 36(2), 111–147 (1974).

25. Kingma, D.P., Ba, J.L.: Adam: A method for stochastic optimization. 1412.6980 (2014).

26. Yeung, N., Summerfield, C.: Metacognition in human decision-making: confidence and error monitoring. Philosophical Transactions of the Royal Society of London, Series B, Biological Sciences 367(1594), 1310–1321 (2012).

27. Mccombe, N., Ding, X., Prasad, G., Gillespie, P., Finn, D.P., Todd, S., Mcclean, P.L., Wong-Lin, K.: Alzheimer’s disease assessments optimized for diagnostic accuracy and administration time. IEEE Journal of Translational Engineering in Health and Medicine 10, 4900809 (2022). doi: 10.1109/JTEHM.2022.3164806.

28. McCombe, N., Liu, S., Ding, X., Prasad, G., Bucholc, M., Finn, D.P., Todd, S., McClean, P.L., Wong-Lin, K.: Practical strategies for extreme missing data imputation in dementia diagnosis. IEEE Journal of Biomedical and Health Informatics 26(2), 818–827 (2022). doi: 10.1109/JBHI.2021.3098511.

29. Xu, Q., Xie, W., Liao, B., Hu, C., Qin, L., Yang, Z., Xiong, H., Lyu, Y., Zhou, Y., Luo, A.: Interpretability of clinical decision support systems based on artificial intelligence from technological and medical perspective: A systematic review. Journal of Healthcare Engineering 2023, 9919269 (2023). doi: 10.1155/2023/9919269.

30. Zanzotto, F.M.: Human-in-the-loop artificial intelligence. Journal of Artificial Intelligence Research 64, 243–252 (2019).

